# Patterns of chemotherapy use and outcomes in advanced non-small-cell lung cancer in relation to age in England: A retrospective analysis of the population-based Systemic Anti-Cancer Treatment (SACT) dataset

**DOI:** 10.1101/2022.10.24.22281434

**Authors:** S. Pilleron, EJA. Morris, D. Dodwell, K.N. Franks

## Abstract

**Objective:** We described the patterns of chemotherapy use and outcomes in patients diagnosed with stage III or IV non-small cell lung cancer (NSCLC) in England.

**Method:** In this retrospective population-based study, we included 20,716 (62% stage IV) patients with NSCLC diagnosed in 2014-17 treated with chemotherapy. We used the Systemic Anti-Cancer Treatment (SACT) dataset to describe changes in treatment plan and estimated 30 and 90-day mortality rates and median, 6- and 12-month overall survival (OS) using Kaplan Meier estimator for patients aged <75 and ≥ 75 by stage. Using flexible hazard regression models we assessed the impact of age, stage, treatment intent (stage III) and performance status on survival.

**Results:** Patients aged ≥ 75 years were less likely to receive 2 or more regimens, more likely to have their treatment modified because of comorbidities and their doses reduced compared to younger patients. However, early mortality rates and overall survival were similar across ages, apart from the oldest patients with stage III disease.

**Conclusion:** This observational study demonstrates that age is associated with treatment choices in an older population with advanced NSCLC in England. Although this reflecting a pre-immunotherapy period, given the median age of NSCLC patients and increasingly older population, these results suggest older patients (>75yrs) may need additional support to access more intense treatments that may influence overall survival.

## Introduction

Non-small cell lung cancer (NSCLC) represents about 85% of new lung cancer diagnoses, affecting about 20,500 people in England annually^1^. Over half of patients with primary NSCLC are diagnosed at an advanced stage ^1^. Net survival is low; in 2010-2014 at 1 year from diagnosis it was estimated at 44% (all stages and ages combined) in England and only 22% at three years ^2^.

Before the routine use of immunotherapy from 2017, cytotoxic chemotherapy was the main treatment for advanced NSCLC management for patients with good performance status ^3^.

As a result of the underrepresentation of older adults in randomized clinical trials ^4^, physicians do not have clear evidence on the benefit-risk balance in this population. Older adults commonly have poorer health status and fitness, more concomitant conditions, age-related physiological changes, and support needs compared to their younger counterparts^5^. Information on outcomes after chemotherapy in older patients with advanced NSCLC and the impact of chemotherapy on mortality at population level is scarce.

The Systemic Anti-Cancer Treatment (SACT) dataset includes data on systemic therapy in all patients diagnosed with cancer in England since 2014 ^6^.

Using the SACT dataset, we described the patterns of chemotherapy use, and associated outcomes in patients diagnosed with stage III or IV NSCLC from 2014 to 2017 in England in relation to age.

## Methods

In this retrospective observational population-based study, we included all patients diagnosed with stage III and IV NSCLC (ICD-10 codes: C34; morphology codes: see supplemental material) between January 1, 2014, and December 31, 2017, aged 18 or over, recorded in the SACT dataset as having received at least one dose of chemotherapy, regardless of treatment intent. We restricted our analysis to the use of cytotoxic chemotherapy for lung cancer (see supplemental Table 1). In case of multiple NSCLC diagnoses in a patient, we kept the first diagnosis in the analysis. All patients were followed for vital status up to December 31, 2018. All cycles of chemotherapy received after this date were excluded from analysis. We further excluded cycles which were recorded as “trial”, were recorded as “not chemo”, “not matched” or “missing treatment group”, and duplicate records (Figure 1). We retrieved characteristics of patients (age, sex, ethnicity) and tumours (morphology, stage at diagnosis) as well as vital status and date of death, if applicable, from the National Cancer Registration and Analysis Service (NCRAS) data linked to SACT data.

**Figure 1.**
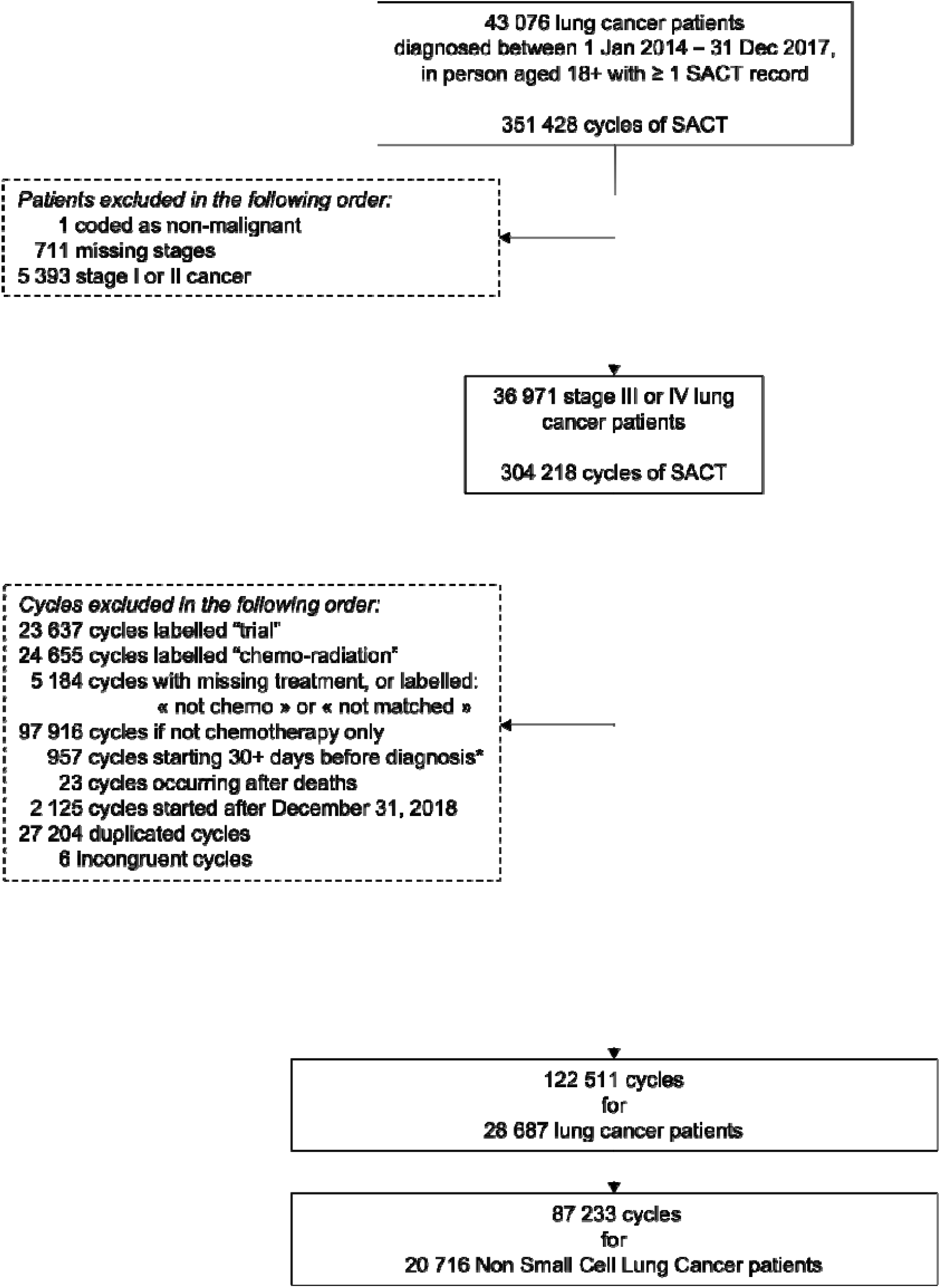
Flow chart

From the SACT dataset, we analysed regimen data that included start date, treatment intent, treatment given, and whether the regimen was adapted based on comorbidities, whether doses were modified or if there were changes in treatment interval. Treatment intent was recoded into three categories: curative (including neo-adjuvant and adjuvant treatment), palliative, or disease modification. A cancer clinician checked for inconsistencies between chemotherapy regimen and treatment intent. In case of inconsistency or missing data, intent was recoded based on review of the regimens used in the context of NSCLC management (supplemental Table 2) and stage. If a chemotherapy regimen may be used in both palliative and curative intent, we kept the original coding. If the original coding was “disease modification”, we kept as is. In stage IV NSCLC, if intent was coded as curative, it was recoded as palliative.

We also analysed the following outcome data ^6^:

- Dose reduction = This identifies any regimens modified by reducing the dose of any anti-cancer drug administered at any point in the regimen after it’s commencement.
- Time delay = This identifies any regimens modified by extending the time between administration dates at any point in the regimen after its commencement.
- Regimen stopped early = This identifies any regimens modified by reducing the administration days below the number originally planned.

For each patient, we kept information regarding the first regimen they received. We also created a variable to indicate whether patients who received a first regimen with curative intent received chemotherapy with palliative intent subsequently.

We also used the start date of cycle of chemotherapy ^6^ and we calculated the number of cycles each patient received.

In our analysis, we kept the performance status recorded at the start of the first chemotherapy cycle and recoded it into 0,1, 2+ and missing. This categorisation was based on NICE guidelines that, at the time, recommended palliative chemotherapy only for patients with a performance score of 0 or 1 ^3^.

### Statistical analysis

All analyses were stratified by stage. Percentages and medians with their interquartile range (IQR) were used to describe categorical and continuous variables (age, number of cycles), respectively.

We estimated mortality rates within 30 and 90 days of the first chemotherapy administration and we used the Kaplan Meier estimator to estimate median, 6- and 12-month overall survival (OS) from the start of the first chemotherapy cycle.

To better describe how age at diagnosis influenced OS, we derived OS from the estimation of individual mortality hazard using a flexible hazard regression model. We modelled the hazard function as the exponential of B-spline of degree 3 with a knot located at the median of the distribution of survival times in patients who died. We included age as a B-spline of degree 3 with a knot at the median of the distribution of age in the whole sample. We first ran models including treatment intent of the first regimen as a factor (stage III), and then performance status as a factor.

Due to the descriptive nature of the study, no statistical tests were performed.

We performed statistical analyses using R statistical software (version 3.4.0; R Development Core Team, 2017) and we used the R ‘mexhaz’ package to model hazard and estimate survival^7^.

### Ethics

North of Scotland Research Ethics Service gave ethical approval for this work (REC/19/NS/0057).

## Results

Out of 43,505 lung cancer patients diagnosed between 1^st^ January 2014 and 31 December 2017, we retained 7,884 stage III and 12,832 stage IV NSCLC (44% females, respectively) (Figure 1).

Age at diagnosis was similarly distributed across stages (Figure 2), with comparable median age (68 [IQR: 62-73] in stage III and 67 [IQR: 60-73] years old in stage IV). Overall, 19.5% and 17.8% patients with stage III and IV NSCLC, respectively, were 75 years or older.

**Figure 2.**
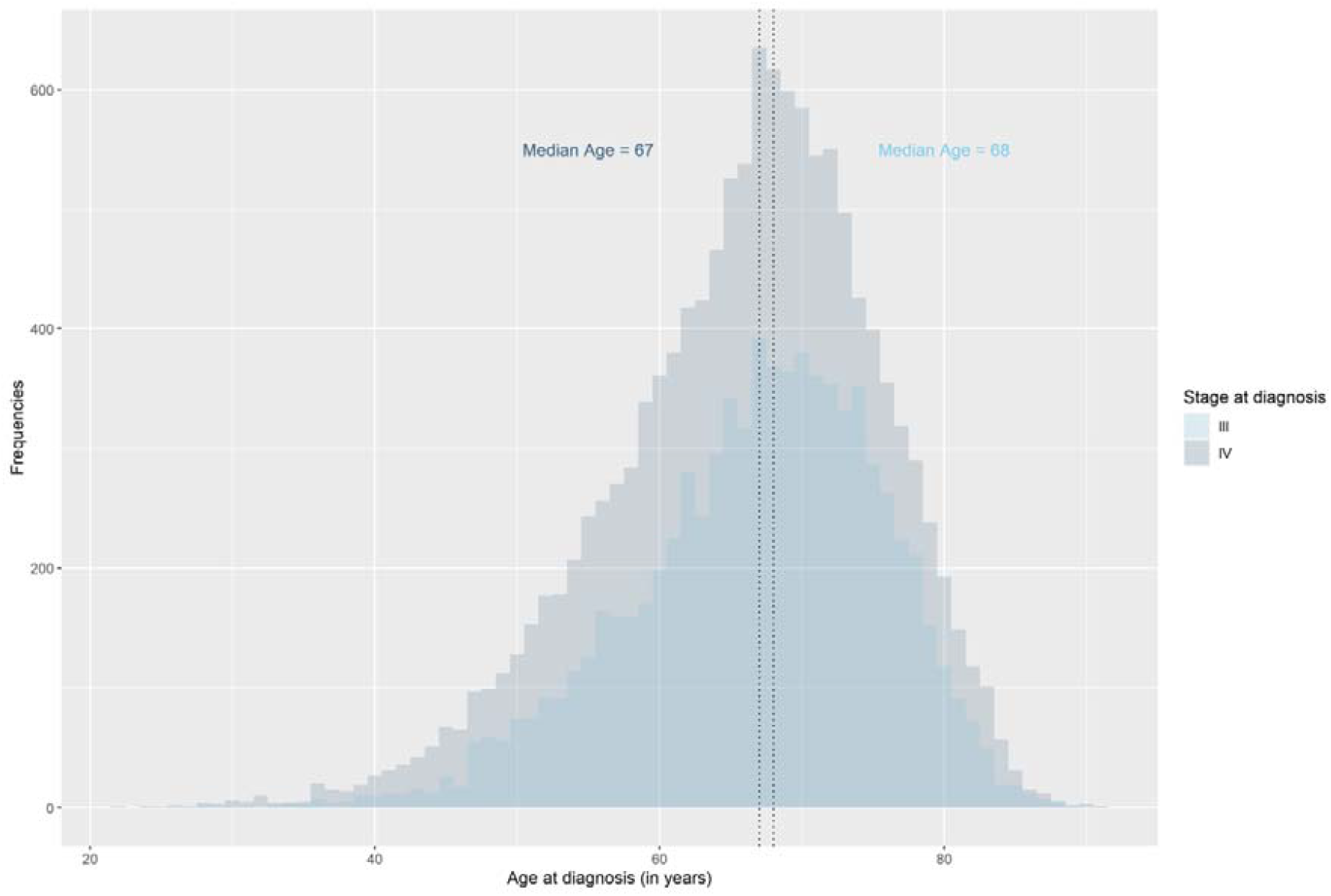
Distribution of age at diagnosis based on stage at diagnosis

Table 1 presents characteristics of patients under and over 75 years old by stage at diagnosis. There were fewer females than males in older than younger patients in both stages (∼38% vs 45%). Ethnicity was equally distributed across age groups in both stages (>90% White). Older patients had poorer performance scores than their younger peers regardless of stage. Older patients with stage III disease were more likely to receive curative treatment than their younger peers (28.6% vs 21.6%, respectively). The median number of cycles received was 4 in all groups except for older patients with stage IV disease (median number = 3).

**Table 1.** Characteristics of patients diagnosed with non-small cell lung cancer in England in 2014-2017 treated with chemotherapy by stage and age group

|  | Stage III |  | Stage IV |  |
| --- | --- | --- | --- | --- |
|  | <75 years old | 75+ years old | <75 years old | 75+ years old |
| n | 6,349 | 1,535 | 10,554 | 2,288 |
| Median age at diagnosis (years [IQR]) | 66.0 [60.0, 70.0] | 77.0 [76.0, 80.0] | 65.0 [58.0, 69.0] | 78.0 [76.0, 80.0] |
| Females - n (%) | 2,856 (45.0) | 589 (38.4) | 4,794 (45.5) | 863 (37.7) |
| Ethnicity - n (%) |  |  |  |  |
| White | 5,962 (93.9) | 1,447 (94.3) | 9,603 (91.1) | 2,161 (94.4) |
| Non-White | 241 (3.8) | 60 (3.9) | 611 (5.8) | 83 (3.6) |
| Unknown/Unstated | 146 (2.3) | 28 (1.8) | 330 (3.1) | 44 (1.9) |
| Performance status at start of the 1st cycle - n (%) |  |  |  |  |
| 0 | 1,645 (25.9) | 320 (20.8) | 2,338 (22.2) | 370 (16.2) |
| 1 | 2,870 (45.2) | 716 (46.6) | 4,647 (44.1) | 1,089 (47.6) |
| 2+ | 540 (8.5) | 196 (12.8) | 1,437 (13.6) | 377 (16.5) |
| Missing | 1,294 (20.4) | 303 (19.7) | 2,122 (20.1) | 452 (19.8) |
| Treatment intent of the 1<sup>st</sup> regimen - n (%) |  |  |  |  |
| Curative | 1,370 (21.6) | 439 (28.6) | - | - |
| Palliative | 4,917 (77.4) | 1,078 (70.2) | 10,477 (99.4) | 2,270 (99.2) |
| Disease modification | 62 (1.0) | 18 (1.2) | 67 (0.6) | 18 (0.8) |
| Change from curative to palliative intent among those who received curative treatment for their 1<sup>st</sup> regimen - n (%) | 838 (61.2) | 244 (55.6) | - | - |
| Median number of cycles recorded [IQR] | 4.00 [2.00, 4.00] | 4.00 [2.00, 4.00] | 4.00 [2.00, 5.00] | 3.00 [2.00, 4.00] |

IQR: Interquartile range

**Table 2.** Changes in chemotherapy treatment plans by stage at diagnosis and age group

|  | Stage III |  | Stage IV |  |
| --- | --- | --- | --- | --- |
|  | <75 | ≥75 | <75 | ≥75 |
| ***First regimen recorded*** |  |  |  |  |
| Adjusted for comorbidity - n (% of non-missing) | 904 (18.6) | 286 (24.3) | 1,494 (18.4) | 432 (24.4) |
| Missing – n (% total) | 1,477 (23.3) | 359 (23.4) | 2,416 (22.9) | 514 (22.5) |
| Dose reduction - n (% of non-missing) | 1,371 (26.9) | 433 (36.1) | 2,347 (27.7) | 648 (35.9) |
| Missing – n (% total) | 1,259 (19.8) | 335 (21.8) | 2,074 (19.7) | 485 (21.2) |
| Time delay - n (% of non-missing) | 1,069 (30.4) | 250 (30.1) | 1,668 (28.3) | 412 (32.4) |
| Missing – n (% total) | 2,837 (44.7) | 704 (45.9) | 4,642 (44.0) | 1,015 (44.4) |
| Stopped earlier - n (% of non-missing) | 882 (18.4) | 215 (19.2) | 1,468 (18.4) | 316 (18.7) |
| Missing – n (% total) | 1,547 (24.4) | 416 (27.1) | 2,580 (24.5) | 600 (26.2) |
| ***In any subsequent regimens recorded*** |  |  |  |  |
| # patients with >1 regimen recorded | 3,119 (49.1) | 681 (44.4) | 5,144 (48.8) | 965 (42.2) |
| Adjustment for comorbidity - n (% of non-missing) | 446 (19.3) | 107 (21.2) | 754 (19.4) | 169 (23.3) |
| Missing – n (% total) | 805 (25.8) | 175 (25.7) | 1,250 (24.3) | 241 (24.9) |
| Dose reduction (% of non-missing) | 832 (33.9) | 226 (43.0) | 1,430 (34.8) | 298 (39.7) |
| Missing – n (% total) | 668 (21.4) | 156 (22.9) | 1,030 (20.0) | 215 (22.3) |
| Time delay - (% of non-missing) | 684 (38.5) | 140 (36.6) | 1,275 (41.1) | 226 (40.0) |
| Missing – n (% total) | 1,341 (43.0) | 298 (43.8) | 2,042 (39.7) | 400 (41.5) |
| Stopped earlier - (% of non-missing) | 467 (20.5) | 86 (18.0) | 914 (23.8) | 151 (21.8) |
| Missing – n (% total) | 839 (26.9) | 204 (30.0) | 1,311 (25.5) | 271 (28.1) |

### Changes in treatment plan

Older patients were more likely to have their treatment adjusted because of the presence of comorbidities (24% vs 18%) and to have reduced doses of chemotherapy than younger patients (36% vs 27%) at both stages of disease.

30% of stage III NSCLC patients had delayed treatment regardless of age. However, treatment delay was more frequently recorded in older stage IV patients compared to younger patients (32% vs 28%). ∼18-19% patients had their treatment stopped earlier than planned regardless of age and stage.

Older NSCLC patients were less likely to have ≥2 regimens reported than younger ones, regardless of stage (42-44% vs 49%). Among patients who received ≥ 1 regimen, the percentage of those for whom a change in treatment plan was recorded was not noticeably different between age groups or stages. Older patients were more likely to have reduced doses of chemotherapy than younger patients (34% in patients under 75 years old in both stages vs 43% and 40% in older patients with stage III and IV, respectively).

*Early mortality and overall survival* (Table 3)

**Table 3.** 30-and 90-day mortality rates, 6-and 12-month and median overall survival from the start of the 1^st^ cycle of chemotherapy in non-small cell lung cancer patients by stage at diagnosis and age group

|  | Stage III |  |  |  | Stage IV |  |  |  |
| --- | --- | --- | --- | --- | --- | --- | --- | --- |
|  | <75 |  | ≥75 |  | <75 |  | ≥75 |  |
|  | # deaths | % [95%CI] | # deaths | % [95%CI] | # deaths | % [95%CI] | # deaths | % [95%CI] |
| 30-day mortality rate | 169 | 2.7 [2.6-2.7] | 48 | 3.1 [3.0-3.2] | 653 | 6.2 [6.1-6.3] | 117 | 5.1 [5.0-5.2] |
| 90-day mortality rate | 533 | 8.4 [8.3-8.5] | 148 | 9.6 [9.4-9.9] | 2,103 | 19.9 [19.8-20.1] | 411 | 18.0 [17.7-18.2] |
| 6-month overall survival | 1,242 | 80.4 [79.4-81.4] | 353 | 77.0 [74.9-79.1] | 4,240 | 59.7 [58.7-60.6] | 903 | 60.4 [58.4-62.5] |
| 12-month overall survival | 2,554 | 59.4 [58.2-60.6] | 721 | 52.8 [50.3-55.4] | 6,989 | 33.2 [32.3-34.1] | 1,514 | 33.4 [31.5-35.4] |
| Median survival (months) | 4,232 | 15.8 [15.2-16.4] | 1,122 | 13.0 [12.2-13.8] | 9,174 | 7.7 [7.5-7.9] | 2,010 | 7.9 [7.5-8.2] |

Note.CI: confidence interval

30-day and 90-day mortality rates were ∼3% and 8-10%, respectively in stage III patients, and 5-6% and 18-20%, respectively, in stage IV patients with no age-related differences within stage.

In respect to overall survival, older patients with stage III disease had a lower 6- and 12-month overall survival by 3.4%-points and 6.6%-points, respectively, and a shortened median survival by 3 months compared to patients ≤75 years of age. However, we did not observe age-related differences in 6- or 12-month OS (60% and 33%, respectively) or median survival (∼8 months in both age groups) in patients with stage IV NSCLC.

Figure 3 shows overall survival over the initial year after the start of the first chemotherapy cycle by age, stage, and treatment intent (Figure 3 A and B), and performance status (Figure 3 C). In stage III NSCLC, overall survival decreased over the first year and as age increased regardless of treatment intent. In stage IV NSCLC, there was no difference in overall survival across ages. There was also no age-related difference in 6- and 12-month overall survival (Figure 3 B) except in stage III patients receiving palliative treatment in whom survival gradually decreased from the age of 65.

**Figure 3.**
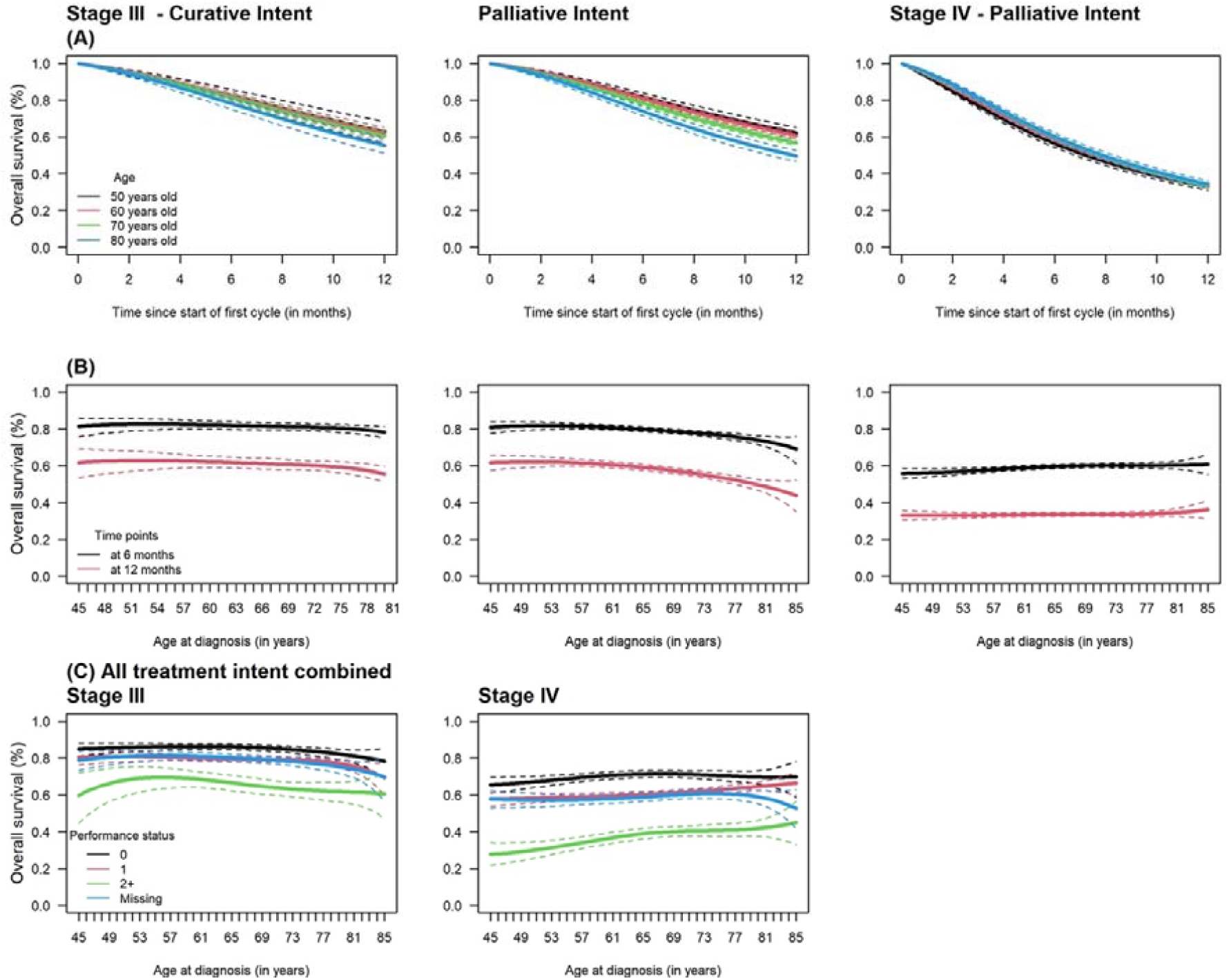
Overall survival (OS) over the initial year of the initiation of chemotherapy at ages 50, 60, 70, and 80; (B) 6 and 12-month OS by age at diagnosis, and (C) 6-month OS by age and performance status in patients diagnosed with stage III and IV non-small cell lung cancer in 2014-2017 and treated with chemotherapy (Note. Solid lines represent overall survival estimates and dashed lines represent 95% confidence intervals)

Finally, 6-month overall survival did not differ between patients with a performance score of 0 or 1 but was poorer in those with score ≥2 in both stage III and IV (Figure 3 C). Overall survival estimates in patients with missing performance status were in between those with score of 0/1 and those with score ≥2.

## Discussion

This population-based study is the first to describe the use of chemotherapy and associated outcomes in a cohort of NSCLC patients treated with chemotherapy in England in relation to age in the era before the introduction of immunotherapy. We showed chemotherapy was more likely to be modified in older patients than their younger peers, and older patients were less likely to have more than one line of treatment. However, age-related differences in early mortality and overall survival were marginal, notably in stage IV NSCLC, suggesting appropriate selection of patients for treatment

There are a few observational studies that describe the use of chemotherapy in NSCLC patients covering more or less the same pre-immunotherapy period. However, most of them were single institution studies and did not stratify by treatment intent or age, making comparisons difficult ^8–12^. Moreover, some estimated survival from diagnosis precluding comparison with our findings ^8^. A study conducted in Leeds in England reported survival estimates stratified by histology and stage and showed an improvement in survival in stage IIIA and III-B-IV NSCLC, mainly in non-squamous cell carcinoma between 2007-2012 and 2013-2017 ^10^. However, results were not provided by age categories or by type of systemic therapy given. A US study reported a median survival of 9.7 months from the initiation of anticancer therapy in stage IV NSCLC patients (median age = 67; 45% females) treated with chemotherapy or immunotherapy (∼20%), higher than in our study, but they did not report estimates by age ^9^.

Although there were more frequent modifications of treatment regimen in older patients, they had similar or somewhat better outcomes than younger patients. This finding suggests the selection of patients to receive chemotherapy was appropriate regardless of age. However, data were not available for the population who did not receive chemotherapy; this precludes any direct comparison. It would seem likely, however, that older patients who received chemotherapy were fitter than those who did not receive.

The main strength of the present study is the inclusion of all patients diagnosed with advanced NSCLC between 2014 and 2017 treated with chemotherapy in England, representing ∼22% of all NSCLC patients diagnosed during the study period (based on an estimate of 82,000 NSCLC patients over the entire period^1^). Other strengths include good completeness of stage at diagnosis (about 2% missing stages), and long follow-up of patients. Our study has, however, limitations. Data (e.g., cycle number or regimen number) may be missing or incorrectly inputted ^6^ and characteristics such as comorbidity, and geriatric conditions (e.g., frailty) for older patients were incomplete precluding in-depth description of those patients who received chemotherapy.

Given these limitations, it would be important to include an objective measure of clinical frailty, in addition to performance status, in future studies focused on older population to better guide which patients are able to safely receive systemic anti-cancer treatment and better tailor the treatment dose and intensity.

Some UK centres having already establish geriatric oncology service eg the Senior Adult Oncology Service at The Christie NHS Foundation NHS Trust, Mancester, England (https://www.christie.nhs.uk/patients-and-visitors/services/senior-adult-oncology-service).

The addition of comprehensive geriatric assessment and a multi-disciiplinary approach with physiotherapy, dietetic, and occupational therapy input may improve the ability to deliver chemotherapy and new therapies (immunotherapy and molecular targeted agents) to this significant population of lung cancer patients.

## Conclusion

This national population-based study provides a snapshot of the chemotherapy use and its associated outcomes in terms of early mortality and overall survival in advanced NSCLC patients under and over 75 years old in England. Our results showed the absence of age-related difference in early mortality and survival. Our findings serve as baseline to monitor chemotherapy outcomes in both age groups as new immunotherapy for NSCLC was made available since 2017.

## Data Availability

De-personalised study data may be made available on request to accredited researchers who submit a proposal that is approved by the PHE Office for Data Release.

## List of abbreviations

NSCLC: Non-small cell lung cancer;
OS: overall survival;
SACT: Systemic Anti-Cancer treatment

## Authors’ contributions

SP designed the study, performed statistical analysis, wrote the first draft. KNF provided clinical expert knowledge.

DD provided clinical input and access to SACT data.

All authors discussed the results and commented on the manuscript.

## Funding information

SP was supported by the European Union’s Horizon 2020 Research and

Innovation Programme, Belgium under the Marie Skłodowska-Curie grant agreement No 842817.

DD is supported by Cancer Research UK (grant no C8225/A21133). Funders had no involvement in the present study.

## Conflict of interest

- SP, EJAM, DD do not have any conflict to report. KNF reports the following:
- Active research funding/grant:
- Avoiding cardiac toxicity in lung cancer patients treated with curative-intent radiotherapy to improve survival. Co-chief investigator (with Prof Faivre-Finn). Funder Yorkshire Cancer Research (YCR)
- PREHABS: Prehabilitation, Radiotherapy Exercise, smoking Habit cessation and Balanced diet Study. Co-chief investigator with Dr Carole Burnett. Funder Yorkshire Cancer Research (YCR)
- CONCOrDE-Study Arm Lead in platform trial of DNA-damage repair inhibitors with radical radiotherapy in stage III NSCLC. (CRUK/AstraZenaca joint funding)
- Yorkshire Lung Screening Trial Biomarker Trial-Co-investigator and PI for Biomarker sub-study of main trial. Funder Yorkshire Cancer Research (YCR)
- UK PI for AstraZeneca PACIFIC-4 Study (currently off license) and CI for AstraZeneca CODAK study

## Honoraria/Advisory Boards

AstraZeneca, Amgen, Boehringer-Ingelheim, Bristol-Meyers-Squibb, Lilly, Roche and Takeda

## Notes

### Competing Interest Statement

SP, EJAM and DD declare no conflict of interest.
KNF reports the following:
Active research funding/grant:
Avoiding cardiac toxicity in lung cancer cases treated with curative-intent radiotherapy to improve survival. Co-chief investigator (with Prof Faivre-Finn). Funder Yorkshire Cancer Research (YCR)
PREHABS: Prehabilitation, Radiotherapy Exercise, smoking Habit cessation and Balanced diet Study. Co-chief investigator with Dr Carole Burnett. Funder Yorkshire Cancer Research (YCR)
CONCOrDE- Study Arm Lead in platform trial of DNA-damage repair inhibitors with radical radiotherapy in stage III NSCLC. (CRUK/AstraZenaca joint funding)
Yorkshire Lung Screening Trial Biomarker Trial- Co-investigator and PI for Biomarker sub-study of main trial. Funder Yorkshire Cancer Research (YCR)
UK PI for AstraZeneca PACIFIC-4 Study (currently off license) and CI for AstraZeneca CODAK study
Honoraria/Advisory Boards: AstraZeneca, Amgen, Boehringer-Ingelheim, Bristol-Meyers-Squibb, Lilly, Roche and Takeda
Educational support: AstraZeneca, Boehringer-Ingelheim, ELEKTA, Pierre Fabre, Novartis, Roche and Takeda.

### Author Declarations

North of Scotland Research Ethics Service gave ethical approval for this work (REC 19 NS 0057).

### Summary of Updates

Following Reviewer's comment and confirmation from KNF, a co-author and clinician, chemotherapy intent for patients with stage IV NSCLC previously coded as "curative" was coded as "palliative". Some results were modified accordingly. One paragraph at the end of the discussion was added. Some minor edits were made.

## References

1. Araghi, M. et al. International differences in lung cancer survival by sex, histological type and stage at diagnosis: an ICBP SURVMARK-2 Study. Thorax (2021) doi:10.1136/thoraxjnl-2020-216555.

2. Araghi, M. et al. Colon and rectal cancer survival in seven high-income countries 2010– 2014: variation by age and stage at diagnosis (the ICBP SURVMARK-2 project). Gut (2020) doi:10.1136/gutjnl-2020-320625.

3. NICE. Lung cancer: diagnosis and management - NICE guideline [NG122]. https://www.nice.org.uk/guidance/ng122 (2021).

4. Sedrak, M. S. et al. Older adult participation in cancer clinical trials: A systematic review of barriers and interventions. CA: A Cancer Journal for Clinicians 71, 78–92 (2021).

5. Mohile, S. G. et al. Practical Assessment and Management of Vulnerabilities in Older Patients Receiving Chemotherapy: ASCO Guideline for Geriatric Oncology. Journal of Clinical Oncology: Official Journal of the American Society of Clinical Oncology 36, 2326–2347 (2018).

6. Bright, C. J. et al. Data Resource Profile: The Systemic Anti-Cancer Therapy (SACT) dataset. International Journal of Epidemiology 49, 15–15l (2020).

7. Charvat, H. & Belot, A. mexhaz: Mixed Effect Excess Hazard Models. R package version 1.5.

8. Soares, M. et al. Real-world treatment patterns and survival outcomes for advanced non-small cell lung cancer in the pre-immunotherapy era in Portugal: a retrospective analysis from the I-O Optimise initiative. BMC Pulm Med 20, 240 (2020).

9. Abernethy, A. P. et al. Real-world first-line treatment and overall survival in non-small cell lung cancer without known EGFR mutations or ALK rearrangements in US community oncology setting. PLOS ONE 12, e0178420 (2017).

10. Snee, M. et al. Treatment patterns and survival outcomes for patients with non-small cell lung cancer in the UK in the preimmunology era: a REAL-Oncology database analysis from the I-O Optimise initiative. BMJ Open 11, e046396 (2021).

11. Vázquez, A. C., Flores, P. G., Ortiz, A. R., Moral, R. G. del & Alcázar-Navarrete, B. Changes in non-small cell lung cancer diagnosis, molecular testing and prognosis 2011– 2016. Journal of Thoracic Disease 10, (2018).

12. Verleye, L. et al. Patterns of care for non-small cell lung cancer patients in Belgium: A population-based study. European Journal of Cancer Care 27, e12747 (2018).

