## Supplemental material 1 for "Patterns of chemotherapy use and outcomes in advanced non-small-cell lung cancer in relation to age in England: A retrospective analysis of the population-based Systemic Anti-Cancer Treatment (SACT) dataset" (from another version of this preprint: version 1, 2022-10-26)

List of morphology codes (ICD10-O2 codes) categorized into non-small cell lung cancer: 8000, 8001, 8003, 8004, 8010, 8012, 8013, 8020, 8021, 8022, 8030, 8031, 8032, 8033, 8034, 8046, 8050, 8052, 8070, 8071, 8072, 8073, 8074, 8075, 8076, 8082, 8083, 8123, 8140, 8141, 8143, 8144, 8145, 8147, 8190, 8200, 8201, 8211, 8230, 8240, 8241, 8243, 8244, 8245, 8246, 8249, 8250, 8251, 8252, 8253, 8255, 8260, 8263, 8310, 8320, 8323, 8370, 8401, 8430, 8440, 8470, 8480, 8481, 8490, 8520, 8550, 8560, 8562, 8570, 8572, 8574, 8575, 8972, 8940, 8980
