## Supplemental Tables for "Patterns of chemotherapy use and outcomes in advanced non-small-cell lung cancer in relation to age in England: A retrospective analysis of the population-based Systemic Anti-Cancer Treatment (SACT) dataset" (from another version of this preprint: version 1, 2022-10-26)

Supplemental Table 1 - Regimen coding

| **Regimen** | **Category** | **Frequencies** |
| --- | --- | --- |
| CARBOPLATIN | Chemotherapy alone | 2375 |
| CARBOPLATIN + DOCETAXEL | Chemotherapy alone | 12 |
| CARBOPLATIN + ETOPOSIDE | Chemotherapy alone | 39339 |
| CARBOPLATIN + GEMCITABINE + PACLITAXEL | Chemotherapy alone | 5 |
| CARBOPLATIN + IRINOTECAN | Chemotherapy alone | 16 |
| CARBOPLATIN + NAB-PACLITAXEL | Chemotherapy alone | 12 |
| CARBOPLATIN + PACLITAXEL | Chemotherapy alone | 1878 |
| CARBOPLATIN + PEMETREXED | Chemotherapy alone | 20721 |
| CARBOPLATIN + TOPOTECAN | Chemotherapy alone | 1 |
| CARBOPLATIN + VINORELBINE | Chemotherapy alone | 10590 |
| CYCLOPHOSPHAMIDE + DOXORUBICIN + VINCRISTINE | Chemotherapy alone | 2080 |
| CISPLATIN | Chemotherapy alone | 132 |
| CISPLATIN + DOCETAXEL | Chemotherapy alone | 47 |
| CISPLATIN + DOXORUBICIN | Chemotherapy alone | 4 |
| CISPLATIN + ETOPOSIDE | Chemotherapy alone | 3897 |
| CISPLATIN + GEMCITABINE | Chemotherapy alone | 1833 |
| CISPLATIN + PACLITAXEL | Chemotherapy alone | 13 |
| CISPLATIN + PEMETREXED | Chemotherapy alone | 15513 |
| CISPLATIN + VINCRISTINE | Chemotherapy alone | 4 |
| CISPLATIN + VINORELBINE | Chemotherapy alone | 7064 |
| DOCETAXEL | Chemotherapy alone | 3909 |
| EC (ETOPOSIDE + PLATINUM (CISPLATINE or CARBOPLATIN)) | Chemotherapy alone | 5 |
| EP (ETOPOSIDE + PLATINUM (CISPLATINE or CARBOPLATIN)) | Chemotherapy alone | 35 |
| ETOPOSIDE | Chemotherapy alone | 83 |
| EVEROLIMUS | Chemotherapy alone | 151 |
| GEMCARBO | Chemotherapy alone | 22079 |
| GEMCITABINE | Chemotherapy alone | 467 |
| PACLITAXEL | Chemotherapy alone | 361 |
| PACLITAXEL + PEMETREXED | Chemotherapy alone | 7 |
| PEMETREXED | Chemotherapy alone | 17931 |
| TOPOTECAN | Chemotherapy alone | 1345 |
| VINORELBINE | Chemotherapy alone | 970 |
| CARBOPLATIN + PEMBROLIZUMAB + PEMETREXED | Chemotherapy + Immunotherapy | 3 |
| ATEZOLIZUMAB + BEVACIZUMAB + CARBOPLATIN + PACLITAXEL | Chemotherapy + Immunotherapy + Targeted Therapy | 31 |
| ATEZOLIZUMAB | Immunotherapy alone | 3714 |
| DURVALUMAB | Immunotherapy alone | 289 |
| IPILIMUMAB | Immunotherapy alone | 2 |
| IPILIMUMAB + NIVOLUMAB | Immunotherapy alone | 7 |
| NIVOLUMAB | Immunotherapy alone | 943 |
| PEMBROLIZUMAB | Immunotherapy alone | 44228 |
| DENOSUMAB | Not chemotherapy - bone | 8581 |
| PAMIDRONATE | Not chemotherapy - bone | 46 |
| ZOLEDRONIC ACID | Not chemotherapy - bone | 2850 |
| ABIRATERONE | Not Lung | 7 |
| ACE | Not Lung | 61 |
| AFLIBERCEPT + FU + IRINOTECAN | Not Lung | 2 |
| BEP | Not Lung | 2 |
| BEVACIZUMAB + IRINOTECAN + MDG | Not Lung | 3 |
| BORTEZOMIB | Not Lung | 4 |
| CABOZANTINIB | Not Lung | 5 |
| CAPECITABINE | Not Lung | 50 |
| CAPECITABINE + CARBOPLATIN | Not Lung | 4 |
| CAPECITABINE + CARBOPLATIN + EPIRUBICIN | Not Lung | 6 |
| CAPECITABINE + CISPLATIN | Not Lung | 8 |
| CAPECITABINE + DOCETAXEL | Not Lung | 1 |
| CAPECITABINE + EPIRUBICIN + OXALIPLATIN | Not Lung | 15 |
| CAPECITABINE + GEMCITABINE | Not Lung | 43 |
| CAPECITABINE + OXALIPLATIN | Not Lung | 15 |
| CAPECITABINE + STREPTOZOCIN | Not Lung | 26 |
| CAPECITABINE + TEMOZOLOMIDE | Not Lung | 91 |
| CAPECITABINE + VINORELBINE | Not Lung | 3 |
| CARBO + FLUOROURACIL | Not Lung | 5 |
| CARBOPLATIN + CETUXIMAB + FLUOROURACIL | Not Lung | 4 |
| CARBOPLATIN + CETUXIMAB + FU | Not Lung | 3 |
| CARBOPLATIN + EPIRUBICIN | Not Lung | 12 |
| CETUXIMAB | Not Lung | 8 |
| CETUXIMAB + CISPLATIN + FU | Not Lung | 2 |
| CHOP | Not Lung | 11 |
| CHOP R | Not Lung | 12 |
| CISPLATIN + FLUORO + STREPTOZOCIN | Not Lung | 3 |
| CISPLATIN + FLUOROURACIL | Not Lung | 5 |
| CTD | Not Lung | 5 |
| CVD | Not Lung | 5 |
| CVP | Not Lung | 1 |
| CVP R | Not Lung | 6 |
| CVP R + GEMCITABINE | Not Lung | 6 |
| CYCLO + DOXORUBICIN + VINCRISTINE | Not Lung | 209 |
| CYCLOPHOSPHAMIDE | Not Lung | 1 |
| CYCLOPHOSPHAMIDE + DOXORUBICIN + ETOPOSIDE | Not Lung | 5 |
| CYCLOPHOSPHAMIDE + ETOPOSIDE | Not Lung | 26 |
| CYTARABINE | Not Lung | 3 |
| DOCETAXEL + PERTUZUMAB + TRASTUZUMAB | Not Lung | 7 |
| DOCETAXEL + TRASTUZUMAB | Not Lung | 2 |
| ECF | Not Lung | 16 |
| ECX | Not Lung | 51 |
| EMA | Not Lung | 1 |
| ENZALUTAMIDE | Not Lung | 3 |
| EOX | Not Lung | 10 |
| EPIRUBICIN | Not Lung | 2 |
| ETOPOSIDE + LOMUSTINE + VINCRISTINE | Not Lung | 17 |
| FCARBOST | Not Lung | 15 |
| FCIST | Not Lung | 1 |
| FEC | Not Lung | 7 |
| FLUOROURACIL | Not Lung | 1 |
| FLUOROURACIL + STREPTOZOCIN | Not Lung | 1 |
| HCX | Not Lung | 5 |
| HORMONES | Not Lung | 115 |
| HYDROXYCARBAMIDE | Not Lung | 3 |
| IMATINIB | Not Lung | 13 |
| IPM | Not Lung | 60 |
| IRINOTECAN | Not Lung | 1 |
| IRINOTECAN + MDG | Not Lung | 29 |
| LANREOTIDE | Not Lung | 198 |
| MCX | Not Lung | 1 |
| METHOTREXATE HIGH DOSE | Not Lung | 3 |
| MITOMYCIN INTRAVESICULAR | Not Lung | 4 |
| MVP | Not Lung | 1 |
| OCTREOTIDE | Not Lung | 93 |
| OXALIPLATIN + MDG | Not Lung | 57 |
| OXALIPLATIN + MDG + PANITUMUMAB | Not Lung | 2 |
| OXALIPLATIN + RALTITREXED | Not Lung | 8 |
| PACLITAXEL + TRASTUZUMAB | Not Lung | 4 |
| PAZOPANIB | Not Lung | 2 |
| PERTUZUMAB + TRASTUZUMAB | Not Lung | 24 |
| RALTITREXED | Not Lung | 1 |
| RITUXIMAB | Not Lung | 14 |
| RUXOLITINIB | Not Lung | 4 |
| STREPTOZOCIN + MDG | Not Lung | 8 |
| TEMOZOLOMIDE | Not Lung | 10 |
| TRASTUZUMAB | Not Lung | 8 |
| VIP | Not Lung | 2 |
| ATEZOLIZUMAB + BEVACIZUMAB | Targeted Therapy + immunotherapy | 3 |
| DOCETAXEL + NINTEDANIB | Targeted Therapy + immunotherapy | 4159 |
| BEVACIZUMAB + CARBOPLATIN + PACLITAXEL | Targeted Therapy + chemotherapy | 2 |
| CARBO + GEFITINIB + PEMETREXED | Targeted Therapy + chemotherapy | 3 |
| AFATINIB | Targeted Therapy alone | 9462 |
| ALECTINIB | Targeted Therapy alone | 549 |
| BEVACIZUMAB | Targeted Therapy alone | 3 |
| BRIGATINIB | Targeted Therapy alone | 301 |
| CERITINIB | Targeted Therapy alone | 1229 |
| CRIZOTINIB | Targeted Therapy alone | 4023 |
| ERLOTINIB | Targeted Therapy alone | 6680 |
| GEFITINIB | Targeted Therapy alone | 7211 |
| LORLATINIB | Targeted Therapy alone | 325 |
| NINTEDANIB | Targeted Therapy alone | 1477 |
| OSIMERTINIB | Targeted Therapy alone | 455 |
| SORAFENIB | Targeted Therapy alone | 18 |
| SUNITINIB | Targeted Therapy alone | 3 |

Supplemental Table 2 – Regimen associated to their treatment intent

| **Chemotherapy regimen** | **Intent** |
| --- | --- |
| CARBOPLATIN | Palliative |
| CARBOPLATIN + DOCETAXEL | Palliative |
| CARBOPLATIN + ETOPOSIDE | Curative |
| CARBOPLATIN + GEMCITABINE + PACLITAXEL | Palliative |
| CARBOPLATIN + NAB-PACLITAXEL | Palliative |
| CARBOPLATIN + PACLITAXEL | Palliative |
| CARBOPLATIN + PEMETREXED | Palliative |
| CARBOPLATIN + VINORELBINE | Curative |
| CAV | Palliative |
| CISPLATIN | Palliative or Curative |
| CISPLATIN + DOCETAXEL | Palliative |
| CISPLATIN + DOXORUBICIN | Palliative |
| CISPLATIN + ETOPOSIDE | Curative |
| CISPLATIN + GEMCITABINE | Palliative or Curative |
| CISPLATIN + PACLITAXEL | Palliative |
| CISPLATIN + PEMETREXED | Palliative |
| CISPLATIN + VINCRISTINE | Palliative |
| CISPLATIN + VINORELBINE | Palliative |
| DOCETAXEL | Palliative |
| EP | Curative |
| ETOPOSIDE | Palliative |
| EVEROLIMUS | Palliative |
| GEMCARBO | Palliative |
| GEMCITABINE | Palliative |
| PACLITAXEL | Palliative |
| PACLITAXEL + PEMETREXED | Palliative |
| PEMETREXED | Palliative |
| TOPOTECAN | Palliative |
| VINORELBINE | Palliative |
